# Statins May Not be Associated with a Reduction in Primary Cardiovascular Events in Patients with Systemic Lupus Erythematosus

**DOI:** 10.64898/2026.06.26.26356369

**Authors:** Lea R. Goren, Michelle Petri, Andrea Fava, Daniel Goldman, Laurence Magder, Luigi Adamo

## Abstract

**Importance:** Cardiovascular disease (CVD) is a leading cause of morbidity and mortality in patients with Systemic Lupus Erythematosus (SLE), due to both traditional CVD risk factors and SLE specific factors. Although statins are first-line therapy for primary prevention of CVD in the general population, it is unclear whether statins protect against first time cardiovascular events (CVEs) in patients with SLE.

**Objective:** Determine whether statins are protective in primary prevention of CVEs among patients with SLE.

**Design, setting, and participants:** This cohort study is a retrospective analysis of a well-characterized, prospective cohort of patients with SLE with patient follow-up beginning in 2013.

**Main outcome and measures:** CVEs were defined as the occurrence of myocardial infarction, thrombotic stroke, onset of angina, or coronary bypass procedure. Statin use in the prior year was quantified based on standardized defined daily doses (DDD). Rates of occurrence were compared using pooled logistic regression. A multivariable model was performed to adjust for possible confounders.

**Results:** The analysis was based on 8708 person-years of follow-up from 1396 cohort participants: 1283 (92%) were women, 567 (41%) Black, and 665 (48%) White. Patients were stratified by use of statin within the last year: none, < standard DDD, or ≥ standard DDD. The rate of events per 1000 person-years was respectively 5.3, 8.5, and 8.0 (p=0.31) within these 3 groups, suggesting potential lack of protective effect of statin treatment. The rates of CVEs among statin versus non-statin users remained the same after adjusting for and stratifying by total cholesterol level (p=0.18). Significantly higher rates of CVEs occurred among those with BMI 25-30 kg/m^2^ (p=0.0066) and those prescribed ≥ 10 mg/day of prednisone (p=0.0003). Multivariable analysis also suggested a potential lack of protective effect of statins against CVEs (OR 1.48; 95% CI, 0.79-2.75; p=0.21883) and diabetes mellitus was found to be independently associated with an increased risk for development of CVEs (OR 4.48; 95% CI 1.99-10.08; p=0.00029).

**Conclusion and Relevance:** Among patients with SLE, statin use may not be protective in primary prevention of CVEs, regardless of statin exposure. Prednisone use, history of diabetes mellitus, and elevated BMI were drivers of increased cardiovascular risk in univariate analysis. Diabetes mellitus persisted as an independent risk factor for CVEs in a multivariable model. Our work reinforces findings from clinical trials which have shown no reduction in subclinical measures of atherosclerosis with statin use among patients with SLE, as well as a mechanistic substudy which demonstrated that statins are ineffective in normalizing the pro-atherogenic changes induced by SLE.

## INTRODUCTION

Cardiovascular disease remains a leading cause of morbidity and mortality in patients with Systemic Lupus Erythematosus (SLE) [1–3]. Despite controlling for traditional cardiovascular risk factors, such as hyperlipidemia and hypertension, the excess risk of experiencing a cardiovascular event (CVE) persists [1,3]. Prior analyses from the Hopkins Lupus Cohort demonstrated a 2.66 increased overall risk of CVEs in patients with SLE as compared to the general population [3]. The excess risk is complex, multifactorial, and incompletely understood, likely a result of underlying immune dysfunction, endothelial injury, and chronic inflammatory changes which promote dysregulated lipid metabolism [4,5] and accelerated atherosclerosis [6,7]. Additional risk can be attributed to other SLE specific factors, such as low C3 levels, presence of lupus anticoagulant, increased global activity scores, and corticosteroid use [1,3,8–10].

In the general population, statins are first-line therapy for the treatment of dyslipidemia and for primary and secondary prevention of atherosclerotic cardiovascular disease [11]. In addition to their role in preventing atherosclerotic progression and decreasing risk of CVEs, statins have demonstrated an immune modulatory benefit independent of their lipid lowering effect [12], including inhibition of proinflammatory mediators, enhancement of endothelial function, and suppression of inflammatory transduction pathways [13–15]. To date however, data supporting the use of statins in the prevention of cardiovascular disease, inhibition of atherosclerosis, and disease activity in patients with SLE is mixed [16–23]. To help address these uncertainties and gain further insight into the utility of statins in primary prevention for patients with SLE, we analyzed statin use and rates of CVEs in the Hopkins Lupus Cohort.

## METHODS

The Hopkins Lupus Cohort is a single-center, longitudinal prospective cohort of patients diagnosed with SLE prior to enrollment in the study. The cohort includes patients enrolled at Johns Hopkins Hospital from 1987 to 2025 and is approved yearly by the Institutional Review Board of the Johns Hopkins University School of Medicine. All patients in the cohort provided written consent prior to entry. At time of entry, patient demographics and laboratory testing relevant to their diagnosis and classification of SLE were collected, including physician global assessment, SLE disease activity index (SLEDAI) [24], organ-specific disease activity, and SLE specific serologies. Patients are followed at least quarterly, or more often if they experience complications or changes in disease activity. Clinical information and laboratory testing are updated at each visit.

The analysis is based on cohort follow-up starting in 2013 when statin use began to be faithfully recorded. Statins were prescribed by either the patient’s primary care provider, cardiologist, or rheumatologist. Statins prescribed include atorvastatin, rosuvastatin, simvastatin, pravastatin, fluvastatin, lovastatin, and pitavastatin. The statin dose at any point of follow-up was based upon the number of days in the previous year that the patient was on a standard dose of statin. Therefore, for the purpose of calculating rates, follow-up for events began after one year of included cohort follow-up. For example, if a patient was in the cohort in January 2013, they were followed for CVEs starting in January 2014. If a patient entered the cohort in January 2015, they were followed for CVEs starting in January 2016.

Statin dose within the last year was defined and calculated using a previously reported method for calculating a defined daily dose (DDD) [16]. For example, if a patient was taking atorvastatin 20 mg/day for the entire prior year, they were classified as ≥ standard DDD. Note that if a patient started taking atorvastatin 20 mg/day within the last year, their follow-up would be classified as < standard DDD until at least one year after initiating statins. CVEs were defined as the occurrence of myocardial infarction, thrombotic stroke, onset of angina, or coronary bypass procedure. Patients with a CVE prior to cohort entry or prior to 2013 were not included in the analysis.

Cohort data were formatted into a dataset which included one record for every person-month of follow-up. Each person-month record included information on whether a patient experienced a CVE during that month of cohort follow-up, as well as an updated medical history of the patient based on information collected at their most recent quarterly visit. Rates of CVEs were compared between patients taking < standard DDD of statin or ≥ standard DDD of statin in the last year. Rates per person-month were converted to rates per 1000 person-years of follow up to assist with interpretation. To assess the relationship between statin dose and CVE, a pooled logistic regression analysis was performed. A multivariable model adjusting for age, diabetes, tobacco use, and prednisone use was also applied to adjust for possible confounders. Statistical analyses were performed using SAS, version 9.4 (SAS Institute).

## RESULTS

The analysis was based on 8708 person-years of follow-up from 1396 different cohort participants of which 1283 (92%) were women, 567 (41%) Black, and 665 (48%) White. Patients were stratified by use of statin within the last year: none, < standard DDD, or ≥ standard DDD. Among patients with no statin use in the prior year, we observed 33 total CVEs over 6276 person-years of follow up, for a rate of 5.3 events per 1000 person-years. Among patients on < standard DDD of statin in the prior year we observed 9 total CVEs (8.5 events per 1000 person-years). Among patients on ≥ standard DDD of statin in the prior year we observed 11 total CVEs (8.0 events per 1000 person-years). The rate of CVEs across the three groups was therefore similar (p=0.31, Table 1). When evaluating rates of MI, we found significantly higher rates of events among patients prescribed ≥ standard DDD of statin (5.7 events per 1000 person-years, p=0.0030). Rate of strokes between statin and non-statin users were not statistically different (p=0.39, Table 1).

**Table 1.**
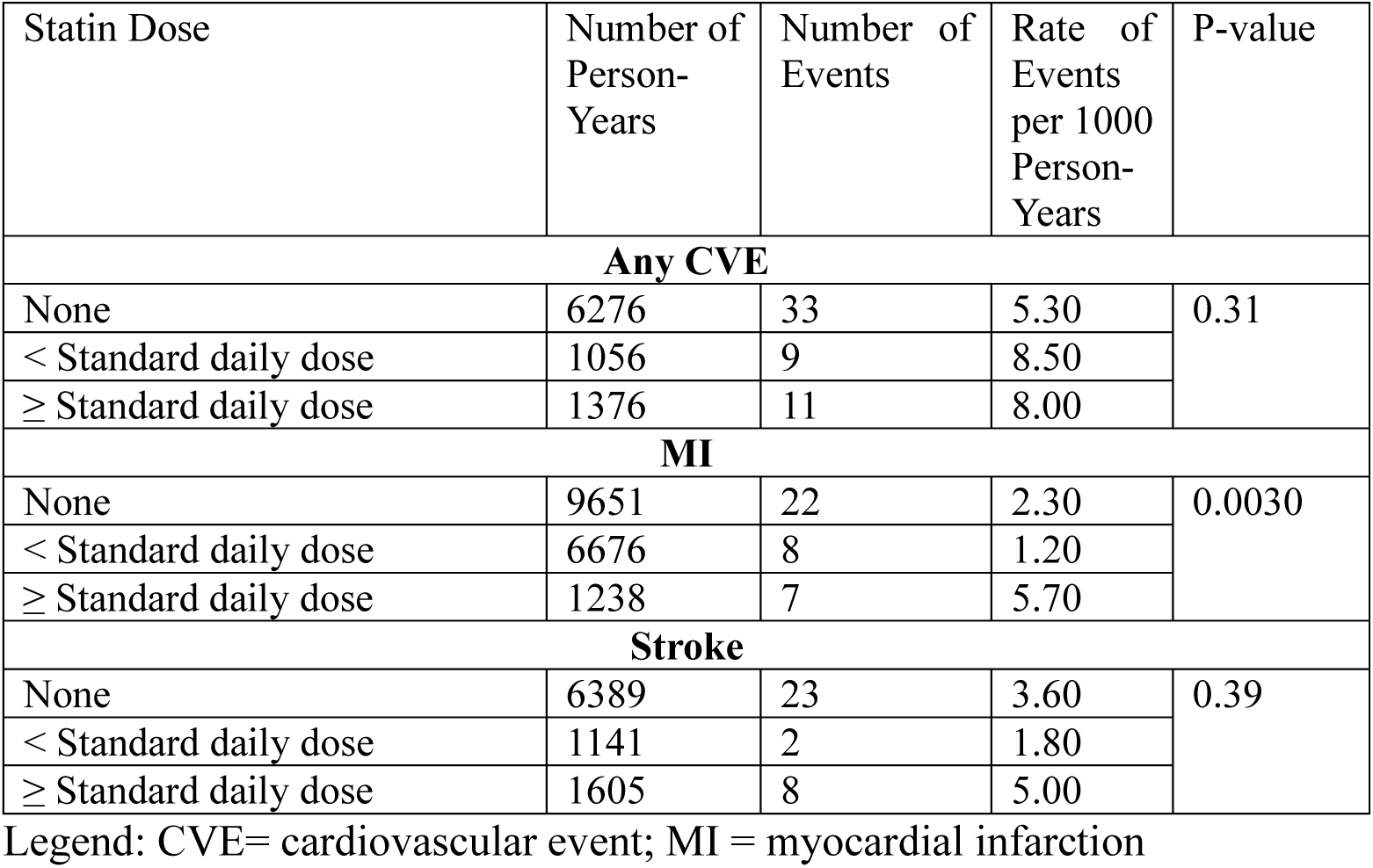
Rates of cardiovascular events in subgroups defined by the mean defined daily dose of statin use in the prior year among all patients with cardiovascular events.

| Statin Dose | Number of Person-Years | Number of Events | Rate of Events per 1000 Person-Years | P-value |
| --- | --- | --- | --- | --- |
| Any CVE |  |  |  |  |
| None | 6276 | 33 | 5.30 | 0.31 |
| < Standard daily dose | 1056 | 9 | 8.50 |  |
| ≥ Standard daily dose | 1376 | 11 | 8.00 |  |
| MI |  |  |  |  |
| None | 9651 | 22 | 2.30 | 0.0030 |
| < Standard daily dose | 6676 | 8 | 1.20 |  |
| ≥ Standard daily dose | 1238 | 7 | 5.70 |  |
| Stroke |  |  |  |  |
| None | 6389 | 23 | 3.60 | 0.39 |
| < Standard daily dose | 1141 | 2 | 1.80 |  |
| ≥ Standard daily dose | 1605 | 8 | 5.00 |  |

Legend: CVE= cardiovascular event; MI = myocardial infarction

To further evaluate the relationship between statin use and CVEs, we examined rates stratified by total cholesterol level (Table 2). No significant differences in rates of total CVEs were found among statin versus non-statin users after adjusting for total cholesterol using logistic regression (p=0.18), with the highest rate of any CVE occurring in patients prescribed statins with total cholesterol levels > 200 mg/dL (10.5 events per 1000 person-years). Patients prescribed statins had significantly higher rates of MI after adjustment for total cholesterol (p=0.0027). No significant differences in rates of stroke were observed after adjusting for total cholesterol (p=0.42).

**Table 2:**
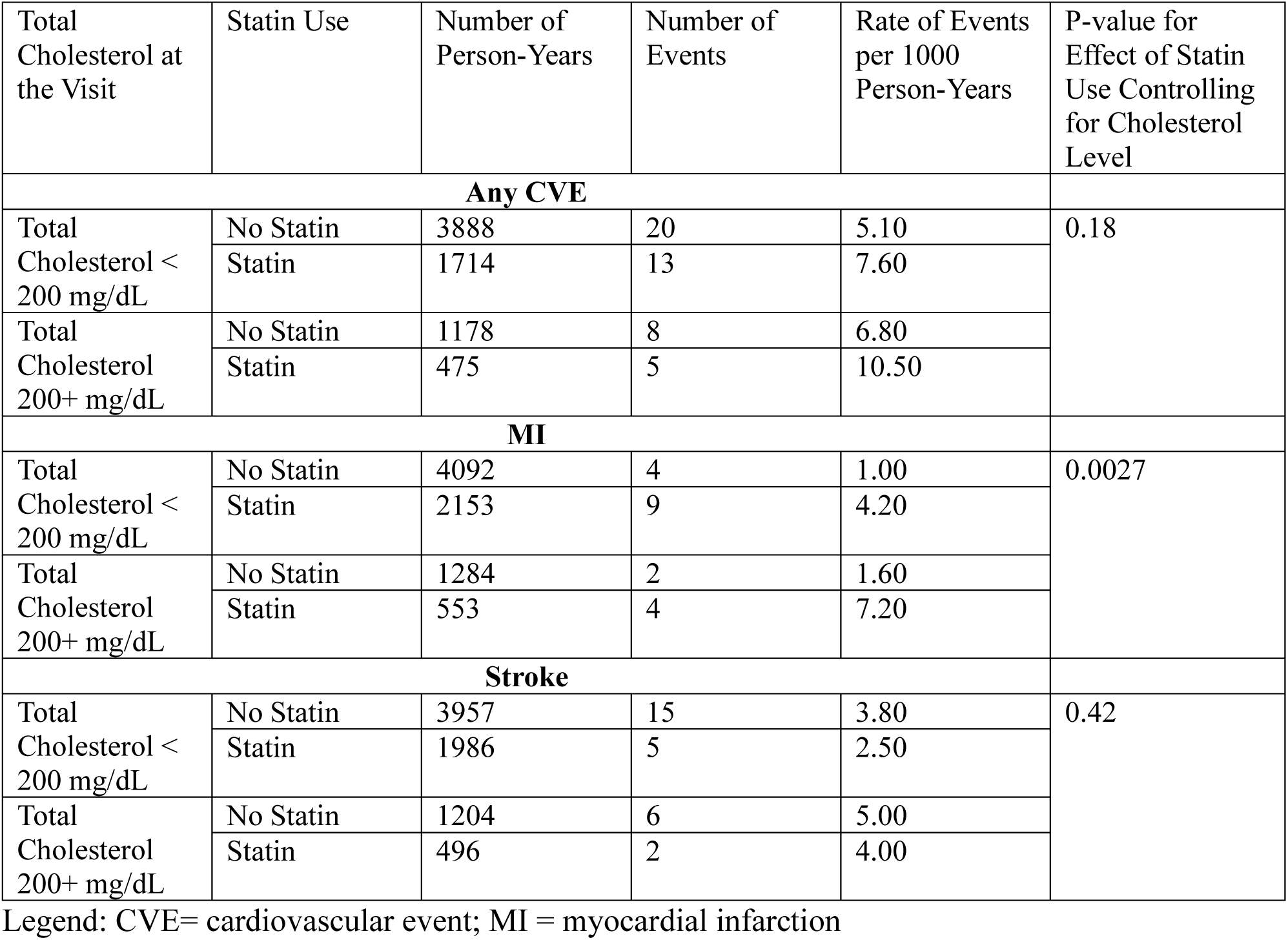
Relationship between statin use and cardiovascular events in strata defined by total cholesterol level.

To determine whether our findings could be a result of confounding by demographic, clinical, serologic, or treatment characteristics, we compared statin users to non-users with respect to covariates (Table 3). Statin users were more likely to be male (11.2% versus 6.0%), over 60 years of age (44.0% versus 15.7%), have history of diabetes mellitus (12.6% versus 5.8%), and have history of tobacco use (33.1% versus 25.1%). Several clinical variables were similar among the two groups, including body mass index (BMI), systolic blood pressure, recent SLEDAI scores, and use of prednisone or hydroxychloroquine.

**Table 3:** Comparison of the follow-up months between non-statin users and statin users with respect to potential confounders.

| Variable | Level | Follow-up Months with No Statins in the Last Year | Follow-up Months with Statins in the Last Year | P-value |
| --- | --- | --- | --- | --- |
| Sex | Female | 5800 (94.0%) | 2158 (88.8%) | 0.0001 |
|  | Male | 370 (6.0%) | 273 (11.2%) |  |
| Age, years | <30 | 557 (9.0%) | 50 (2.0%) | <0.0001 |
|  | 30-39 | 1593 (25.8%) | 179 (7.4%) |  |
|  | 40-49 | 1669 (27.1%) | 382 (15.7%) |  |
|  | 50-59 | 1382 (22.4%) | 752 (30.9%) |  |
|  | 60+ | 966 (15.7%) | 1069 (44.0%) |  |
| Race | White | 2849 (46.2%) | 1302 (53.6%) | 0.036 |
|  | Black | 2682 (43.5%) | 944 (38.8%) |  |
|  | Other | 639 (10.4%) | 185 (7.6%) |  |
| Body Mass Index, kg/m<sup>2</sup> | <25 | 2205 (41.7%) | 754 (33.6%) | 0.13 |
|  | 25-30 | 1437 (27.2%) | 646 (28.8%) |  |
|  | 30-35 | 804 (15.2%) | 477 (21.3%) |  |
|  | 35+ | 838 (15.9%) | 368 (16.4%) |  |
| Systolic Blood Pressure, mmHg | <120 | 2304 (43.7%) | 663 (29.6%) | 0.19 |
|  | 120-129 | 1562 (29.6%) | 726 (32.4%) |  |
|  | 130-139 | 892 (16.9%) | 518 (23.1%) |  |
|  | 140+ | 516 (9.8%) | 333 (14.9%) |  |
| History of Diabetes Mellitus | No | 5811 (94.2%) | 2126 (87.4%) | <0.0001 |
|  | Yes | 358 (5.8%) | 305 (12.6%) |  |
| Ever Smoked | No | 4617 (74.9%) | 1626 (66.9%) | 0.0005 |
|  | Yes | 1550 (25.1%) | 803 (33.1%) |  |
| Years since SLE diagnosis | <5 | 762 (12.1%) | 126 (5.2%) | 0.0010 |
|  | 5-15 | 2256 (36.0%) | 607 (25.0%) |  |
|  | 15+ | 3258 (51.9%) | 1698 (69.8%) |  |
| Recent SLE Disease Activity Index Score | <3 | 3495 (65.3%) | 1612 (71.1%) | 0.40 |
|  | 3+ | 1854 (34.7%) | 656 (28.9%) |  |
| History of anti-dsDNA | No | 2362 (37.6%) | 951 (39.1%) | 0.11 |
|  | Yes | 3914 (62.4%) | 1481 (60.9%) |  |
| History of nephrotic-level proteinuria | No | 5206 (83.0%) | 1856 (76.3%) | 0.063 |
|  | Yes | 1070 (17.0%) | 575 (23.7%) |  |
| History of neurologic involvement | No | 5799 (92.4%) | 2258 (92.9%) | 0.12 |
|  | Yes | 477 (7.6%) | 173 (7.1%) |  |
| History of antiphospholipid antibodies | No | 2672 (42.6%) | 896 (36.9%) | 0.69 |
|  | Yes | 3604 (57.4%) | 1535 (63.1%) |  |
| Prednisone use | No | 4381 (80.0%) | 1789 (78.6%) | 0.90 |
|  | <10 mg/day | 870 (15.9%) | 416 (18.3%) |  |
|  | ≥ 10 mg/day | 228 (4.2%) | 71 (3.1%) |  |
| Plaquenil Use | No | 675 (12.4%) | 417 (18.5%) | 0.39 |
|  | Yes | 4787 (87.6%) | 1831 (81.5%) |  |

Legend: SLE = Systemic Lupus Erythematosus; anti-dsDNA = anti-double stranded DNA

We also investigated the relationship between patient characteristics and rate of CVEs among all patients in the analysis (Table 4). Significantly higher rates of CVEs occurred among patients with BMI 25-30 kg/m^2^ (10.6 events per 1000 person-years, p=0.0066) and among those taking ≥ 10 mg/day of prednisone (23.4 events per 1000 person-years, p=0.0003).

**Table 4:** Rate of cardiovascular events by various predictors.

| Variable | Level | Number of Person Years | Number of Events | Rate of Events per 1000 Person-Years |  |
| --- | --- | --- | --- | --- | --- |
| Age, years | <50 | 4430 | 22 | 5.00 | 0.69 |
|  | 50-59 | 2134 | 19 | 8.90 |  |
|  | 60+ | 2035 | 12 | 5.90 |  |
| Sex | Female | 7957 | 48 | 6.00 | 0.59 |
|  | Male | 643 | 5 | 7.80 |  |
| Race | White | 4151 | 24 | 5.80 | 0.87 |
|  | Black | 3625 | 23 | 6.30 |  |
|  | Other | 824 | 6 | 7.30 |  |
| Body Mass Index, kg/m<sup>2</sup> | <25 | 2959 | 9 | 3.00 | 0.0066 |
|  | 25-30 | 2083 | 22 | 10.60 |  |
|  | 30-35 | 1281 | 11 | 8.60 |  |
|  | 35+ | 1206 | 6 | 5.00 |  |
| Systolic Blood Pressure, mmHg | <120 | 2967 | 17 | 5.70 | 0.23 |
|  | 120-129 | 2289 | 13 | 5.70 |  |
|  | 130-139 | 1411 | 8 | 5.70 |  |
|  | 140+ | 849 | 10 | 11.80 |  |
| Diabetes Mellitus | No | 7937 | 47 | 5.90 | 0.32 |
|  | Yes | 663 | 6 | 9.00 |  |
| Smoker | No | 6243 | 37 | 5.90 | 0.65 |
|  | Yes | 2353 | 16 | 6.80 |  |
| Years since SLE diagnosis | <5 | 887 | 6 | 6.80 | 0.54 |
|  | 5-14 | 2864 | 14 | 4.90 |  |
|  | 15+ | 4956 | 33 | 6.70 |  |
| Recent SLE Disease Activity Index Score | <3 | 5108 | 27 | 5.30 | 0.075 |
|  | 3+ | 2510 | 22 | 8.80 |  |
| History of anti-dsDNA | No | 3313 | 13 | 3.90 | 0.055 |
|  | Yes | 5395 | 40 | 7.40 |  |
| History of nephrotic-level proteinuria | No | 7062 | 45 | 6.40 | 0.46 |
|  | Yes | 1645 | 8 | 4.90 |  |
| History of neurologic involvement | No | 8058 | 47 | 5.80 | 0.30 |
|  | Yes | 650 | 6 | 9.20 |  |
| History of antiphospholipid antibodies | No | 3568 | 26 | 7.30 | 0.19 |
|  | Yes | 5139 | 27 | 5.30 |  |
| Prednisone use | None | 6171 | 31 | 5.00 | 0.0003 |
|  | <10 mg/day | 1285 | 10 | 7.80 |  |
|  | ≥ 10 mg/day | 299 | 7 | 23.40 |  |
| Plaquenil Use | No | 1092 | 7 | 6.40 | 0.93 |
|  | Yes | 6618 | 41 | 6.20 |  |

Legend: SLE = Systemic Lupus Erythematosus; anti-dsDNA = anti-double stranded DNA

Based on the findings in Tables 3 and 4, a multivariable logistic regression model was fit to adjust for age, history of diabetes, tobacco use, and prednisone use (Table 5). After adjusting for potential confounders, multivariable analysis suggested a potential lack of protective effect of statins against CVEs (OR 1.48; 95% CI, 0.79-2.75; p=0.21883). However, the presence of diabetes was independently associated with an increased risk for the development of CVEs (OR 4.48; 95% CI 1.99-10.08; p=0.00029).

**Table 5:** Multivariable model for cardiovascular events among patients taking statins.

| Variable | Comparison | Odds Ratio (95% CI) | P-value |
| --- | --- | --- | --- |
| Statin Use | Yes vs. No | 1.48 (0.79, 2.75) | 0.21883 |
| Sex | Male vs. Female | 1.23 (0.48, 3.13) | 0.66100 |
| Age | 50-60 years vs <50 years | 1.85 (0.94, 3.63) | 0.07616 |
|  | 60+ years vs <50 years | 1.09 (0.49, 2.43) | 0.83430 |
| Diabetes | Yes vs. No | 4.48 (1.99,10.08) | 0.00029 |
| Smoker | Yes vs. No | 0.92 (0.33, 2.60) | 0.87830 |
| Prednisone | 10+mg/d vs. less | 1.05 (0.56, 1.95) | 0.88829 |

## DISCUSSION

In this study of a large, well-defined longitudinal cohort of patients with SLE and no prior CVE, we found that statin use may not be associated with a protective effect against CVEs, regardless of statin exposure. We observed no significant differences in rates of total CVEs or stroke among statin and non-statin users, while rates of MI were significantly higher among patients prescribed ≥ standard DDD of statin. After controlling for total cholesterol levels, we observed no differences in rates of total CVEs or stroke in statin vs non-statin users. Notably, rates of MI were highest among statin users, regardless of total cholesterol level. Although total number of CVEs were small in our cohort, our findings help expand our understanding of the impact of statin use in patients with SLE and suggest that statins may not be an effective therapy for primary prevention of CVEs in this unique population.

The mechanisms involved in accelerated atherosclerosis in patients with SLE are multifactorial, reflecting the complex interplay between traditional cardiovascular risk factors, such as obesity and diabetes, as well as SLE specific risk factors-including inflammatory signaling pathways, complement activation, and altered lipid profiles. To date, randomized trials have not provided consistent evidence for the use of statins in cardiovascular disease prevention among patients with SLE. For example, the Lupus Atherosclerosis Prevention Study (LAPS), a randomized trial investigating the impact of atorvastatin versus placebo on atherosclerosis progression in 200 patients with SLE, did not find significant differences in coronary artery calcium, carotid plaque, or carotid intima media thickness over the two year study duration [19]. In contrast, a randomized trial by Plazak et al. compared atorvastatin to placebo in 60 patients with SLE and demonstrated a significant increase in coronary calcium among the placebo group after one year [17]. Other statins, such as rosuvastatin, have also been studied in clinical trials. Mok et al. described significant improvement in inflammatory markers and low density lipoprotein cholesterol (LDL-C) levels among 72 patients with SLE and subclinical atherosclerosis taking low-dose rosuvastatin after one year of therapy as compared to placebo [25]. Reductions in carotid intima-media thickness were also noted after 24 months between the groups, though the findings were nonsignificant. Although statin use has been shown to improve lipid profiles in patients with SLE, outcomes in atherosclerotic progression are not consistent among these trials.

Importantly, studies evaluating the impact of statins on CVEs in patients with SLE are limited and the data presented above do not necessarily support a protective effect of statins in terms of reduction of CVEs. In fact, while in SLE patients statin use was associated with inhibition of coronary calcium progression in one study [17], in the general population, initiation of statin therapy is associated with an increase in coronary calcium which is interpreted as a sign of plaque stabilization that correlates with a reduction of cardiovascular events [26]. To date, one randomized trial showed that fluvastatin reduced the risk of major cardiac events by 73.4% after 7-8 years of follow-up among patients with SLE who underwent renal transplant; however, the findings were non-significant (p=0.064) [22] and a population of renal transplant recipients is not representative of most SLE patients. An observational cohort study of 4095 patients with SLE and hyperlipidemia by Yu et al. demonstrated a significant reduction in all-cause mortality, coronary artery disease, cerebrovascular disease, and end-stage renal disease among patients with SLE taking high dose statins. [16]. However, a systematic review comparing these studies found no effect of statins in the prevention of CVEs [27], consistent with findings from our cohort.

Although statins have known immunomodulatory benefits, their impact in reducing inflammatory markers are mixed. For example, while meta-analyses have demonstrated significant reductions in plasma C-reactive protein among patients with SLE treated with statins [21,23], atorvastatin failed to decrease levels of high sensitivity C-reactive protein or reduce disease activity in the LAPS trial [19]. These findings were further investigated on a cellular level in a mechanistic substudy, which showed that statins were unable to normalize the pro-atherogenic changes induced by SLE and were not effective in altering cholesterol transport gene expression in human macrophages from patients in the LAPS trial [28]. Our findings reinforce the notion that statin exposure might not be protective against first time CVEs and are consistent with findings from the LAPS trial and cellular investigations.

Among all patients included in our cohort, we found a significant increase in rate of total CVEs in patients taking prednisone in our univariate analysis, with doses ≥ 10 mg/day resulting in highest risk. Corticosteroid use is a known risk factor for cardiovascular events among patients with SLE [10] and a dose-dependent increase in CVE rates has previously been described in the Hopkins Lupus Cohort [3]. However, in our multivariable analysis among patients taking statins, we did not find corticosteroid use to be independently associated with increased rates of CVEs, which may reflect a specific protective effect of statin on steroid induced CVEs or limited statistical power.

We also observed a significant increase in rates of CVEs among patients with elevated BMI, particularly in those with BMI 25-30 kg/m^2^ and 30-35 kg/m^2^. Elevated BMI is a key modifiable risk factor in the development of cardiovascular disease among the general population; however, the impact of BMI on cardiovascular outcomes in patients with SLE is heterogenous and overall unclear. For example, in a cohort of 49 patients with SLE, BMI > 25 kg/m^2^ was independently associated with increased internal carotid artery wall thickness and the risk of carotid atherosclerosis increased 16% for each kg/m^2^ of BMI increase [29]. In contrast, in a cohort of 61 patients with SLE, low BMI was associated with progression of carotid artery intima-media thickness [30]. Our findings suggest that higher BMI is associated with increased rates of CVEs and that it should therefore be addressed as a modifiable risk factor in this patient population. Additional studies are needed to further refine this observation.

Among patients in our cohort, those with diabetes were at highest risk for the development of CVEs, especially among statin users. Diabetes is a well-known risk factor for the development of cardiovascular disease and in our multivariable analysis, we found that diabetes is independently associated with an increased risk of CVE by more than four-fold (OR 4.48; 95% CI 1.99-10.08; p=0.00029). This is consistent with analyses from the RELESSER study, which demonstrated a greater than two-fold risk of CVE among patients with SLE and diabetes [31]. These findings suggest that aggressive diabetes control is paramount in reducing cardiovascular risk in this unique population and that this increased risk is not effectively addressed by statin therapy.

Our study has the notable strength of being focused on a large, well characterized longitudinal cohort of SLE patients. However, it has several limitations that should be kept in mind when considering our findings. First, numbers of total CVEs in the cohort were low and LDL-C data was not available for all patients. Second, patients in the cohort were from a single center, which may impact the generalizability of our results. Third, we assumed patients took statins as prescribed; however, they were followed up at least every three months, with medication reconciliations completed at all visits. Fourth, criteria for statin initiation were not standardized; statin therapy was started according to the discretion of the medical team. This is a strength to the extent that allowed us to produce “real world data,” but also a weakness because we cannot exclude unmeasurable heterogeneity between statin and non-statin users and cannot exclude confounding by indication.

## Conclusion

Among patients with SLE, statin use may not be protective in primary prevention of CVEs, regardless of statin exposure. Prednisone use, history of diabetes mellitus, and elevated BMI were drivers of increased cardiovascular risk in univariate analysis. The presence of diabetes persisted as an independent risk factor for CVEs in a multivariable model focused on statin users. To date, the ideal strategy for preventing CVEs in patients with SLE remains unclear. Our findings suggest that the complex inflammatory and proatherogenic mechanisms which drive atherosclerosis in SLE might not be mitigated by statins and call for additional multicenter studies to define appropriate strategies to reduce cardiovascular disease burdens in patients with SLE. Since the increased burden of CVEs in SLE patients is likely immune mediated, immunomodulatory therapies with well-defined effects on CVE burden, such as proprotein convertase subtillsin/Kexin Type 9 inhibitors, colchicine, and interleukin-1 blockade [32–34], appear promising candidates for such studies.

## Data Availability

All data produced in the present study are available upon reasonable request to the authors.

## Author contributions

Study concept and design: LA, MP, LRG. Acquisition, analysis, and interpretation of data: MP, LM, DG, AF, LRG, LA. Writing-original draft: LRG. Writing-review & editing: LRG, MP, AF, DG, LM, LA.

